# Advanced Spatio-Temporal Neural Networks for Malaria Prevalence Forecasting in Sub-Saharan Africa

**DOI:** 10.1101/2025.11.12.25340054

**Authors:** Vikalp Srivastava, Manoj Kumar, Sanika Vaidya, Abdessamad Tridane

**Affiliations:** School of Engineering and Science, Indian Institute of Technology Madras, Zanzibar; Department of Mathematics Institute of Chemical Technology, Mumbai, Department of Mathematical Sciences, United Arab Emirates University

## Abstract

Accurate malaria prevalence forecasting is critical for timely interventions in Sub-Saharan Africa, where the disease remains a major health burden. Traditional regression models often struggle to capture the complex interactions between the epidemiological, environmental, and spatial factors that drive malaria transmission. This study develops and compares three distinct deep learning approaches for predicting the Plasmodium falciparum parasite rate in children (PfPR_2−10_), using historical prevalence data from 1900 to 2015 and climate variables such as temperature and precipitation sourced from NASA’s POWER project. The first framework is a Spatiotemporal Transformer developed to capture intricate local dependencies between covariates. The second is a Patch-based Time Series Transformer (PatchTST) designed for robust point forecasting across temporal dimensions. The third approach employs a Spatio-Temporal Neural Ordinary Differential Equation (Neural ODE) model that learns the underlying continuous-time dynamics governing malaria prevalence, enabling smooth temporal interpolation and extrapolation. All three models were further extended with uncertainty estimation using Monte Carlo Dropout, providing probabilistic forecasts in addition to point predictions. Our findings highlight a key architectural trade-off: for this dataset, characterized by short time series and strong cross-feature interactions, the Spatiotemporal Transformer yielded the highest predictive accuracy. Conversely, while the Neural ODE demonstrated slightly lower point-forecast accuracy, it effectively captured temporal continuity and uncertainty dynamics, offering deeper insights into malaria’s spatio-temporal evolution. This comparative analysis provides a nuanced perspective, offering decision-makers a choice between highly accurate models for direct forecasting and probabilistic frameworks for risk assessment and strategic planning.

## 1 Introduction

Malaria remains one of the most persistent and devastating public health challenges in Sub-Saharan Africa, affecting millions each year and placing a substantial burden on healthcare systems, economies, and communities. According to the World Health Organization’s latest reports [9], the region accounts for approximately 95% of global malaria cases and 96% of malaria deaths, with children under five years of age being disproportionately affected. A key aspect of effective malaria control lies in the ability to accurately forecast disease prevalence across different temporal and spatial scales [6]. Such forecasting capabilities enable policymakers, health organizations, and international aid agencies to allocate resources more efficiently, deploy preventative measures proactively, and respond to potential outbreaks before they reach critical levels. However, malaria transmission is influenced by a complex and dynamic interplay of epidemiological, environmental [12], and spatial factors, making accurate prediction a challenging and multifaceted task [5].

Environmental factors such as temperature and precipitation significantly influence mosquito breeding cycles and survival rates [7]. Spatial factors including population density, urbanization levels, access to healthcare, and economic development impact both disease transmission patterns and treatment outcomes. Additionally, biological factors such as vector species distribution, parasite drug resistance patterns, and host immunity levels add further complexity to the prediction challenge.

Traditional regression models often fall short in capturing these complex interactions between the epidemiological, environmental, and spatial factors that drive malaria transmission due to their reliance on linear assumptions and limited capacity to model complex interactions between variables [1–3]. Early machine learning approaches, while more flexible, typically treat spatial and temporal dimensions as independent features, failing to capture the inherent spatio-temporal correlations that characterize vector-borne diseases like malaria.

Recent advances in machine learning and deep learning have shown significant promise in infectious disease modeling and diagnosis. [11] explored interpretable ML frameworks for malaria prediction, addressing the crucial need for transparency in health applications. Complementing this, [8] developed an ensemble AI-enabled MIoT system for automated malaria parasite detection, while [10] introduced MalNet-DAF, a dual-attentive fusion deep model that enhances classification performance. Similarly, [4] demonstrated the utility of YOLO-mp for real-time malaria parasite detection and counting, advancing automated diagnostics. Beyond malaria, [14] applied deep epidemiological modeling with black-box knowledge distillation for accurate COVID-19 predictions, highlighting the broader applicability of AI in epidemiology.

This study develops and compares three distinct deep learning approaches for predicting the *Plasmodium falciparum* parasite rate among children aged 2 to 10 (PfPR_2−10_), using historical prevalence data (1900–2015) and climate variables—temperature and precipitation—sourced from NASA’s POWER project. The first framework is a Spatiotemporal Transformer developed to capture intricate, local dependencies between covariates. The second is a Patch-based Time Series Transformer (PatchTST) designed for robust point forecasting across temporal dimensions. The third approach employs a Spatio-Temporal Neural Ordinary Differential Equation (Neural ODE) model that learns the underlying continuous-time dynamics governing malaria prevalence, enabling smooth temporal interpolation and extrapolation. All three models were further extended with uncertainty estimation using Monte Carlo Dropout, providing probabilistic forecasts in addition to point predictions.

## 2 Methodology

### 2.1 Problem Formulation and Mathematical Framework

Our malaria forecasting problem can be formally defined as a supervised spatio-temporal regression task where we aim to predict future malaria prevalence given historical multi-modal data. Let 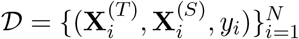 represent our dataset of *N* samples, where:

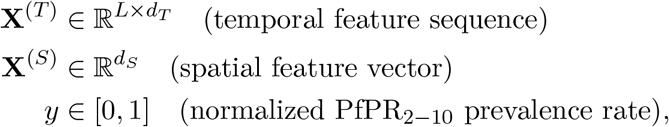

where *L* represents the sequence length, *d*_*T*_ denotes the total number of time-varying features (including epidemiological indicators, derived patterns, and climate variables), and *d*_*S*_ represents the static spatial dimensions (geographic coordinates and location-specific statistics).

Our objective is to learn a function 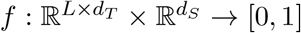 such that:

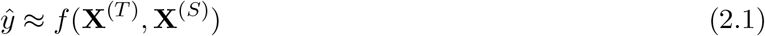

The learning objective minimizes the expected prediction error:

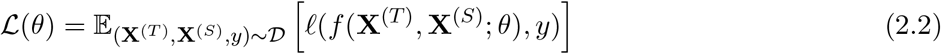

where *ℓ* (·, ·) is the loss function, *θ* represents the learnable parameters, and the expectation is taken over the data distribution.

### 2.2 Dataset Description and Characteristics

Our prediction framework is built by combining two primary sources of information: a comprehensive database of historical malaria cases and a detailed record of environmental data.

- **Malaria Prevalence Data:** The foundation of our study is the **Malaria SSR Database** [13], which provides 28,632 geo-referenced malaria measurements from surveys conducted across Sub-Saharan Africa. Each record contains crucial details, including **temporal features** (like the year, month, and the age range of people surveyed) and **spatial features** (such as the survey’s coordinates and administrative boundaries). We also enriched this data with engineered features to capture seasonal trends and short-term historical patterns.
- **Climate Data:** To provide environmental context, we used monthly meteorological data from **NASA’s POWER database**. This dataset includes key climate variables, such as **temperature** (T2M) and **precipitation** (PRECTOTCORR), at a 0.5^*°*^ × 0.625^*°*^ resolution. We also engineered more advanced climate features, such as interactions and ratios between variables. To combine these datasets, we aligned each malaria record with its corresponding climate data using a nearest-neighbor approach, effectively matching each survey to its closest weather station data.

Our **target variable** for prediction is the **PfPR**_2−10_. This is the standard metric for malaria prevalence, representing the percentage of *Plasmodium falciparum* infection found in children between the ages of 2 and 10.

### 2.3 Data Pre-processing and Feature Engineering

Our data pipeline transforms raw epidemiological surveillance data into a feature-rich format through systematic imputation, engineering, and scaling procedures.

a. **Missing Value Imputation**
  i. **Age bounds (LoAge, UpAge):** Imputed with defaults (2, 10).*These represent the lower and upper age bounds of surveyed populations, typically reflecting the 2-10 year age group commonly used in malaria prevalence surveys*.
  ii. **Survey metrics (Ex, Pf):** Imputed using location-specific median values. *Ex represents the number of individuals examined in a survey, while Pf represents the number testing positive for Plasmodium falciparum. Location-specific imputation preserves local epidemiological patterns*.
b. **Feature Engineering** We constructed four categories of engineered features:
  1. **Temporal Features:** To capture time-dependent patterns and seasonality.
    i. **Year normalization:** Scales survey year (YY) to [0,1] range for numerical stability.

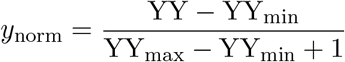

*Prevents arbitrary magnitude differences between years from biasing the model*.
    ii. **Cyclical month encoding:** Represents survey month (MM) as circular features.

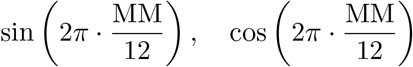

*Captures the cyclical nature of seasons, ensuring December and January are treated as adjacent months rather than distant values*.
    iii. **Lagged features:** Incorporates historical values from previous time steps.

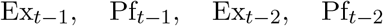

*Provides temporal context by including examination counts and positive cases from one and two time steps prior, capturing short-term transmission dynamics*.
    iv. **Moving averages (3-point):** Smooths time series to reduce measurement noise.

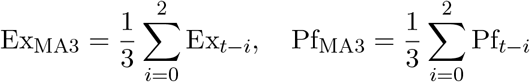

*Reduces short-term fluctuations in surveillance data, revealing underlying epidemiological trends*.
  2. **Derived Epidemiological Features:** To quantify disease-specific metrics.
    i. **Age range:** Width of the surveyed age cohort.

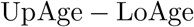

*Accounts for heterogeneity in survey design, as different age ranges exhibit varying malaria susceptibility patterns*.
    ii. **Prevalence rate:** Normalized proportion of positive cases.

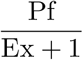

*Computes the infection rate relative to survey size, providing a standardized measure independent of sample size variations*.
  3. **Derived Spatial Features:** To encode geographic characteristics.
    i. **Latitude-longitude interaction:** Captures unique geographic positioning.

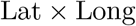

*Creates a composite spatial feature representing the interaction between latitude and longitude coordinates from the survey location*.
    ii. **Distance from equator:** Absolute latitude as a malaria risk indicator.

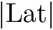

*Proximity to the equator correlates strongly with malaria endemicity due to favorable climatic conditions for Anopheles mosquito breeding*.
    iii. **Hemisphere indicator:** Binary encoding of north/south hemisphere.

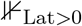

*Distinguishes Northern (1) from Southern (0) hemisphere locations, accounting for opposing seasonal patterns in malaria transmission*.
  4. **Location-Specific Aggregates:** To characterize endemic patterns.
    i. **Historical means:** Average surveillance metrics per location.

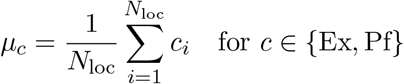

*Captures baseline endemicity levels, distinguishing high-transmission hotspots from low-risk areas based on historical survey data*.
    ii. **Historical standard deviations:** Temporal variability per location.

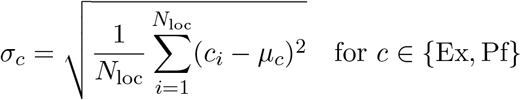

*Quantifies transmission stability, differentiating stable endemic regions from areas prone to epidemic outbreaks*.
c. **Sequence Construction**
  i. **Sliding windows** of length *L* = 6 observations were generated per location. *Creates sequential input patterns enabling the model to learn temporal dependencies and seasonal transmission cycles from consecutive survey observations*.
d. **Categorical Encoding**
  i. **Label encoding** converts country identifiers (COUNTRY) to integers.

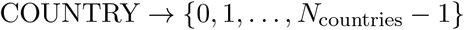

*Transforms categorical country labels from the AFR ADMIN2 Code classification into numerical representations suitable for neural network processing*.
e. **Target Smoothing**
  i. A **3-point moving average** was applied to the target variable (PfPR2-10).

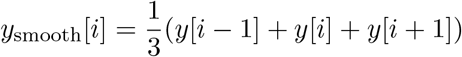

*Reduces measurement noise in parasite prevalence rates, which can exhibit high variance due to small sample sizes in individual surveys*.
f. **Normalization and Scaling**
  i. **Feature standardization (Z-score):** Centers and scales all input features.

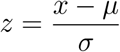 *Ensures all features contribute equally to model training regardless of their original measurement scales, preventing dominance by high-magnitude variables*.
  ii. **Target scaling (Min-Max):** Normalizes PfPR2-10 prevalence rates to [0,1].

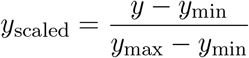 *Aligns target variable range with the sigmoid activation function output, facilitating stable gradient-based optimization during training*.

### 2.4 Incorporating Climate Data

We formulate malaria prevalence prediction as a supervised spatiotemporal forecasting problem. The model is provided a sequence of time-varying features **X**^(*T*)^ and a corresponding static spatial feature vector **X**^(*S*)^. The objective is to predict the malaria prevalence *y*.

The temporal feature sequence 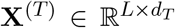 contains all time-varying inputs, including epidemiological features (e.g., month, age, survey data), derived patterns (e.g., lags, moving averages), and raw climate variables (e.g., monthly temperature and precipitation). The static spatial context is represented by 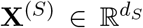, which encompasses geographic coordinates and location-specific statistics, such as the long-term mean and standard deviation of the climate variables for a particular location.

The prediction task aims to learn a mapping function 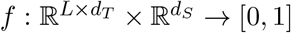 such that:

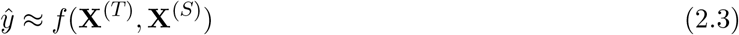

This formulation allows the model to leverage both short-term temporal patterns and long-term spatial and environmental contexts for improved forecasting accuracy.

### 2.5 Enhanced Spatio-Temporal Transformer (ESTT) Architecture

Our proposed architecture (shown in Figure 1 (a)) extends standard transformer encoders with specialized components designed for spatiotemporal climate-disease modeling. The model consists of four primary modules: temporal encoding, transformer processing, spatial integration, and fusion-based prediction.

**Figure 1:**
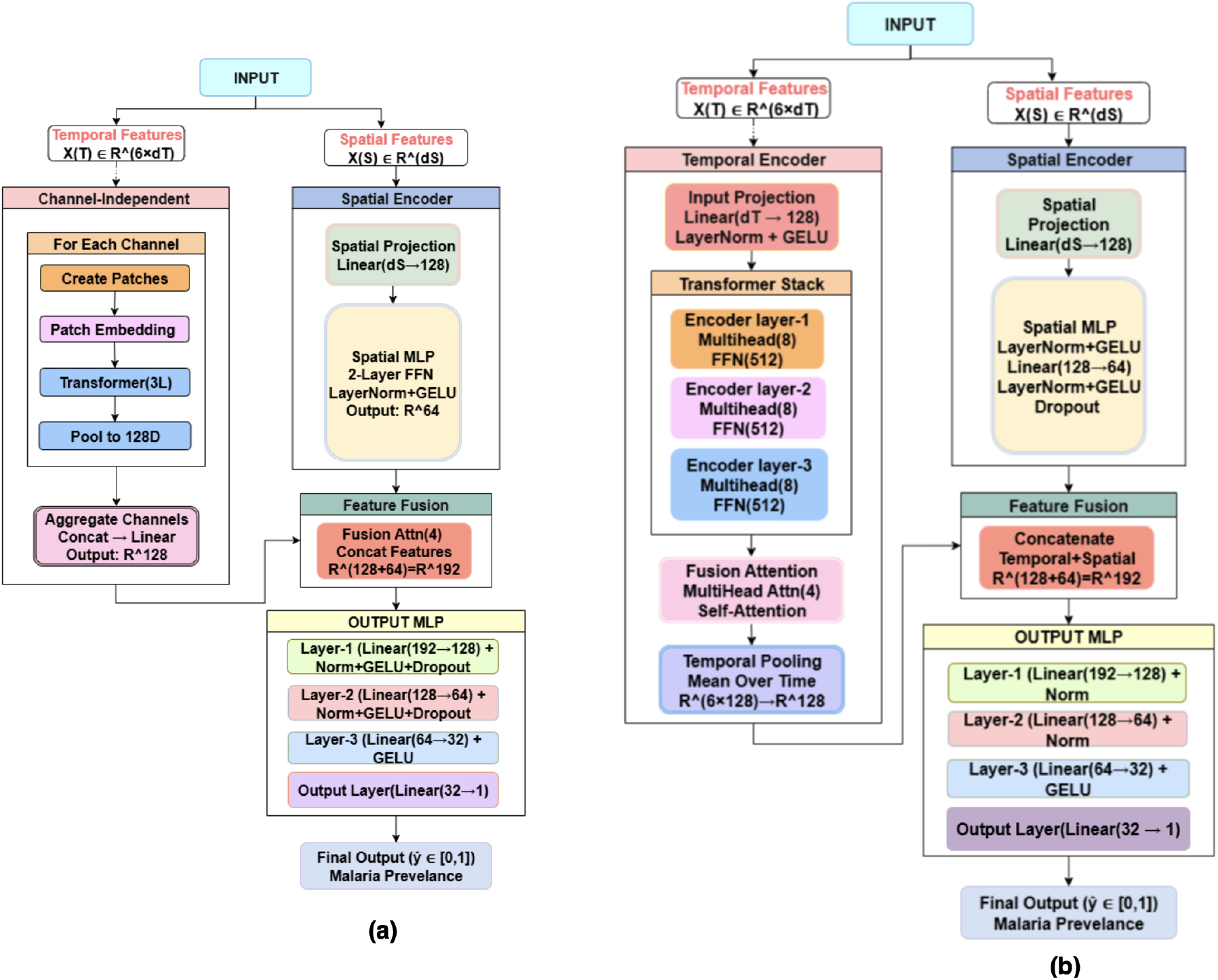
**(a)** PatchTST Architecture for Climate-Disease Modeling. **(b)** Enhanced Spatiotemporal Transformer Architecture for Climate-Disease Modeling.

#### Temporal Feature Projection

The input temporal sequence 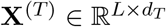 (where *L* = 6 for sequence length) undergoes linear projection to the model’s hidden dimension, followed by layer normalization and nonlinear activation:

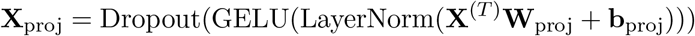

where 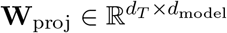 and 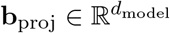 are learnable parameters.

#### Positional Encoding

To incorporate temporal ordering information, we add learnable positional embeddings to the projected features:

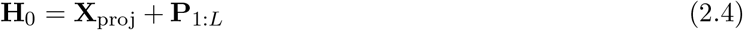

where 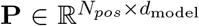 represents the full matrix of position embeddings (*N*_*pos*_ = 100) and **P**_1:*L*_ denotes the first *L* embeddings corresponding to the sequence length.

#### Multi-Layer Transformer Encoding

The temporally encoded features pass through 3 transformer encoder layers with a **norm-first** architecture and residual connections:

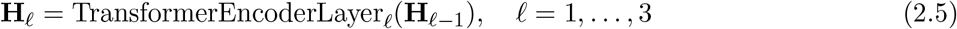

Each transformer layer incorporates multi-head self-attention (*n*_heads_ = 8) with GELU activations and increased feedforward dimensions (4 × *d*_model_) to capture complex temporal dependencies.

#### Enhanced Spatial Feature Processing

Spatial features 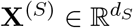 (including geographic coordinates, location statistics, and derived spatial interactions) undergo projection and multi-layer processing:

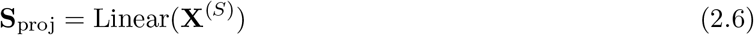

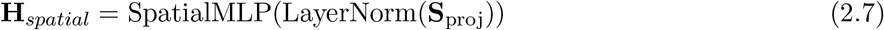

The spatial MLP consists of two fully connected layers with layer normalization, GELU activation, and dropout, reducing the feature dimensionality to *d*_model_*/*2.

#### Attention-Based Temporal-Spatial Fusion

We employ a multi-head self-attention layer (*n*_heads_ = 4) to refine the temporal representations before pooling:

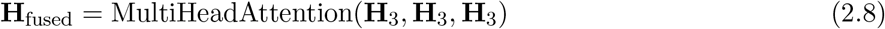

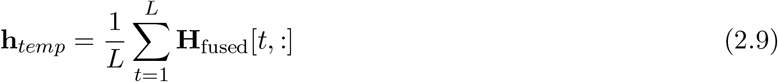

#### Enhanced Prediction Module

The final prediction is generated by a deep MLP that takes the concatenated temporal and spatial feature vectors:

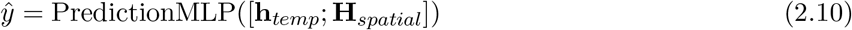

The prediction MLP incorporates four layers with dimensions (*d*_model_ + *d*_model_*/*2) → *d*_model_ → *d*_model_*/*2 → *d*_model_*/*4 → 1, utilizing layer normalization, GELU activation, and dropout for robust training. More details can be found in Appendix A.

## 3 PatchTST-Based Spatiotemporal Forecasting Architecture

We implement an enhanced PatchTST architecture (shown in Figure 1 (b)) specifically adapted for spatiotemporal malaria forecasting, leveraging patch-based decomposition to efficiently process heterogeneous epidemiological and climatic time series data.

The key innovation of our approach lies in decomposing time series into patches and processing each feature channel independently, enabling specialized learning for different data modalities (epidemiological vs. climatic variables) while maintaining computational efficiency.

### 3.1 Patch-Based Decomposition

Given the multivariate time series 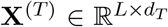, the model first separates it into *d*_*T*_ channels. For each channel 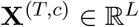, patches of length *P* and stride *S* are extracted:

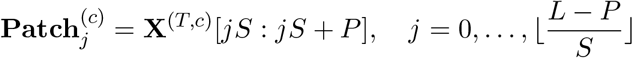

Each patch is flattened, linearly projected to 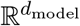, and added with learnable positional encodings.

### 3.2 Channel-Independent Processing

Each feature channel **X**^(*T,c*)^ ∈ ℝ^*L*^ is patch-embedded and passed through a dedicated Transformer encoder with multi-head self-attention:

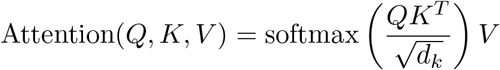

Outputs from all *d*_*T*_ channels are concatenated and linearly projected to form the aggregated temporal representation **h**_agg_.

### 3.3 Spatial Feature Integration

Spatial attributes 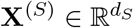 (e.g., coordinates, climate statistics) are projected via:

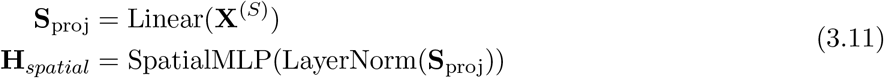

The SpatialMLP uses GELU activation and dropout.

### 3.4 Spatiotemporal Fusion and Prediction

The aggregated temporal representation **h**_agg_ is first refined using a self-attention mechanism to capture the most salient temporal patterns. The resulting features, **h**_*temp*_, are then fused with the processed spatial features **H**_*spatial*_ via concatenation.

1. **Temporal Feature Refinement (Self-Attention):**

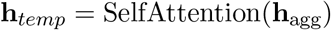
2. **Spatiotemporal Fusion (Concatenation) and Final Prediction:**

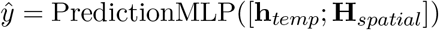

### 3.5 Training

Smooth L1 loss with L2 regularization:

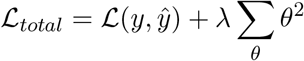

Optimization uses AdamW with cosine annealing warm restarts and gradient clipping for stability.

This novel adaptation of PatchTST for spatiotemporal malaria forecasting represents a significant advancement in epidemiological modeling, combining the efficiency of patch-based processing with the sophistication of attention mechanisms specifically tailored for climate-enhanced disease prediction in resource-constrained settings. More details can be found in Appendix A.

## 4 Neural ODE-Based Spatiotemporal Malaria Forecasting

We implement a Neural ODE architecture (shown in Figure 2) specifically adapted for spatiotemporal malaria forecasting, leveraging continuous-time dynamics to model disease progression patterns while incorporating both epidemiological and climatic variables. The key innovation lies in modeling temporal evolution as a continuous dynamical system, enabling the model to naturally handle irregular temporal sampling and capture smooth disease progression dynamics.

**Figure 2:**
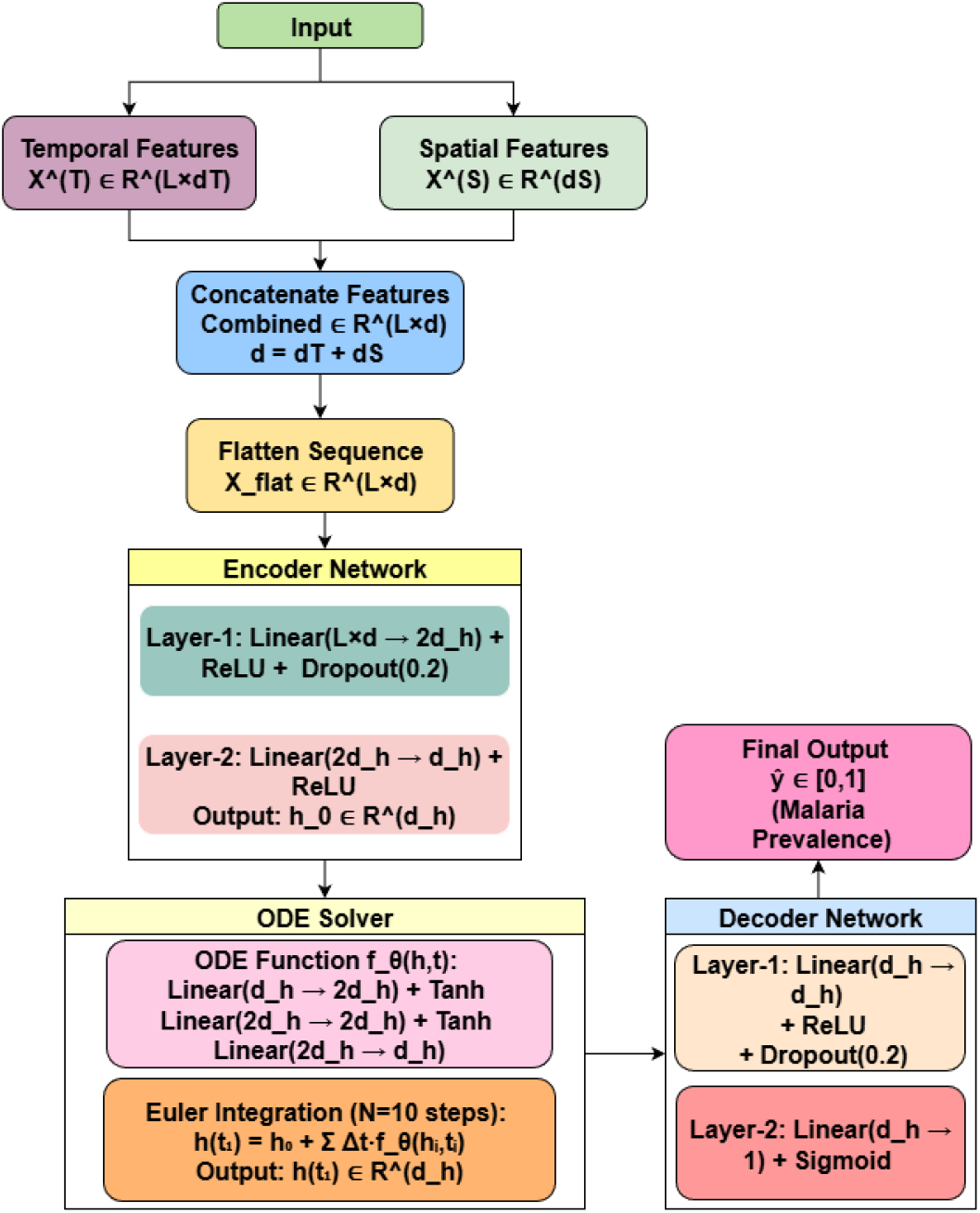
Neural ODE Architecture for Climate-Disease Modeling.

### 4.1 Encoder Network

The encoder maps the flattened sequence input **x**_flat_ ∈ ℝ^*L·d*^ (where *L* is sequence length and *d* is feature dimension) to a hidden state 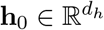 :

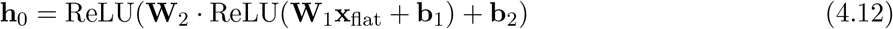

with dropout (*p* = 0.2) applied between layers for regularization.

### 4.2 ODE Dynamics Function

The temporal evolution of the hidden state is modeled as a continuous dynamical system:

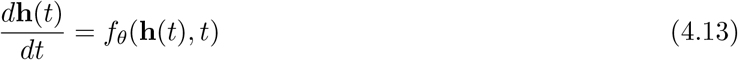

where *f*_*θ*_ is parameterized by a three-layer neural network with tanh activations:

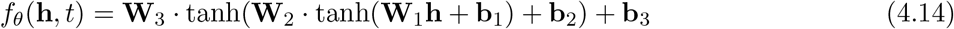

### 4.3 Numerical ODE Solver

The initial value problem is solved using forward Euler method with *N* = 10 integration steps:

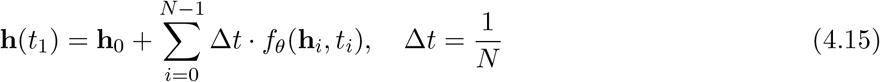

### 4.4 Decoder Network

The evolved hidden state is decoded to the final prediction:

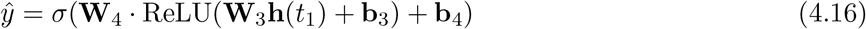

where *σ* is the sigmoid function ensuring *ŷ* ∈ [0, 1].

This Neural ODE approach provides a continuous-time framework for malaria forecasting that naturally handles irregular temporal sampling and captures smooth disease progression dynamics while integrating heterogeneous spatiotemporal features.

## 5 Experimental Results

In this section, we have conducted a comprehensive comparative analysis of three statistical regression models (shown in Figure 3), traditional machine learning approaches, and advanced deep learning architectures including spatiotemporal transformers, PatchTST, and Neural ODE models. These models predict malaria parasite prevalence (‘Pf’) based on geographical coordinates (‘Lat’ and ‘Long’). Table 1 presents a comprehensive performance comparison between different models.

**Table 1:** Comparison of traditional and proposed models.

| Methodology | RMSE | MAE | $R^2$ |
| --- | --- | --- | --- |
| **Spatiotemporal Neural ODE** | **9.680** | **6.650** | **0.844** |
| **Spatiotemporal Transformer** | **9.688** | **6.497** | **0.841** |
| **PatchTST** | **17.448** | **12.535** | **0.486** |
| Poisson Regression | 124.4111 | 41.6535 | 0.0065 |
| Binomial Regression | 146.2883 | 38.7158 | -0.3736 |
| Linear Regression | 124.3484 | 41.6155 | 0.0075 |

**Figure 3:**
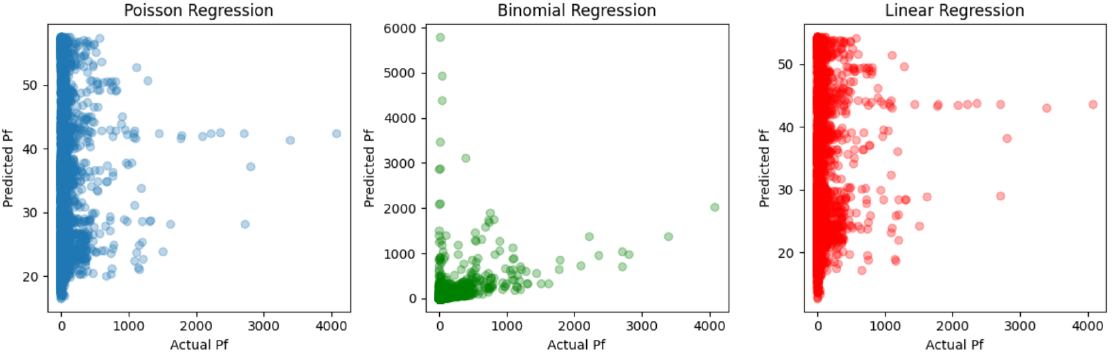
Actual vs. Predicted values for the Poisson, Binomial, and Linear Regression models.

### 5.1 Multi-Country Evaluation

We also evaluate our models across five sub-Saharan African countries with diverse malaria transmission patterns and geographic characteristics: Congo, Tanzania, Nigeria, Mozambique, and Uganda. Each country presents unique epidemiological challenges, ranging from high-burden equatorial regions to seasonal transmission zones, enabling comprehensive assessment of model generalizability across different architectures.

### 5.2 Performance Analysis

The results demonstrate varying performance patterns across different model architectures and geographic regions, providing insights into the effectiveness of climate integration and model complexity for malaria prediction.

#### Neural ODE Analysis

The Neural ODE results shown in Table 2 reveal an unexpected pattern. The baseline model, without climate data, actually outperforms the climate-enhanced variant across most countries. On average, the baseline model achieves a 3.2% lower MAE (9.473 vs. 9.778), a 4.6% higher *R*^2^ (0.734 vs. 0.702), and a 5.5% lower RMSE (11.719 vs. 12.396).This counterintuitive finding is especially stark in Nigeria, where the baseline model’s MAE is 20.8% better and its *R*^2^ is 14.2% higher. This suggests that the continuous-time dynamics modeled by Neural ODEs might already be capturing the implicit, smooth patterns of seasonality driven by climate, making the explicit climate features redundant or even just adding noise.In fact, the baseline model performed better in four of the five countries (Nigeria, Congo, Uganda, and Tanzania). Mozambique was the only exception, showing a very slight improvement with climate integration (MAE 9.510 vs. 9.613), indicating that the benefit of this extra data is not guaranteed and varies by region.

**Table 2:**
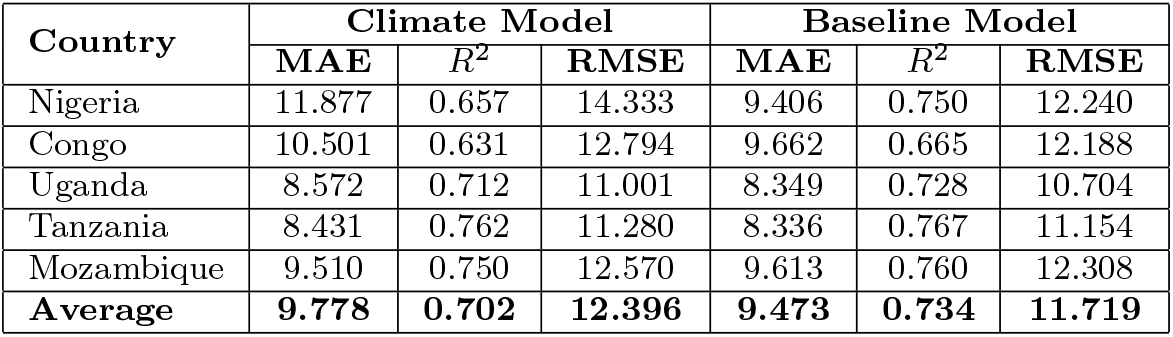
Performance comparison of Neural ODE models (Climate vs. Baseline) for malaria prediction across different countries.

#### Machine Learning Models Analysis (Table 3)

Traditional ensemble methods demonstrate highly variable responses to climate integration. Random Forest consistently shows marginal performance variations across countries, with improvements ranging from +0.4% to +1.2% MAE in most regions (Tanzania, Nigeria, Mozambique, Congo), while Uganda exhibits slight degradation (-0.9%). This stability suggests Random Forest’s robustness to additional features but limited sensitivity to climate variables, likely due to its tendency to distribute importance across many features. Gradient Boosting, however, reveals striking heterogeneity: while Tanzania and Nigeria show modest improvements (+0.3% to +0.6%), Congo experiences severe degradation (-8.8% MAE increase) with climate features. Mozambique (-2.0%) and Uganda (-0.7%) also saw their performance worsen, though more moderately. This pronounced variability indicates that Gradient Boosting’s sequential learning process may be highly sensitive to feature quality and regional data characteristics. The Congo anomaly suggests potential overfitting to noisy climate data or misalignment between climate variables and local transmission dynamics.Overall, ensemble methods achieve competitive performance (*R*^2^ ranging 0.626-0.805) but show inconsistent climate benefits, highlighting the importance of feature selection and region-specific tuning for traditional ML approaches.

**Table 3:** Performance comparison of Random Forest and Gradient Boosting models for malaria prediction across different countries.

| Country | Model | Climate |  | Baseline |  | MAE Impr. (%) |
| --- | --- | --- | --- | --- | --- | --- |
| | | MAE | $R^2$ | MAE | $R^2$ | |
| Uganda | Random Forest | 8.524 | 0.727 | 8.448 | 0.734 | -0.9 |
|  | Gradient Boosting | 7.849 | 0.757 | 7.797 | 0.759 | -0.7 |
| Congo | Random Forest | 10.358 | 0.626 | 10.480 | 0.626 | +1.2 |
|  | Gradient Boosting | 9.035 | 0.654 | 8.302 | 0.704 | -8.8 |
| Tanzania | Random Forest | 7.792 | 0.788 | 7.828 | 0.782 | +0.5 |
|  | Gradient Boosting | 7.436 | 0.802 | 7.480 | 0.805 | +0.6 |
| Nigeria | Random Forest | 9.594 | 0.760 | 9.677 | 0.763 | +0.8 |
|  | Gradient Boosting | 9.269 | 0.768 | 9.301 | 0.763 | +0.3 |
| Mozambique | Random Forest | 9.497 | 0.761 | 9.539 | 0.760 | +0.4 |
|  | Gradient Boosting | 9.242 | 0.778 | 9.060 | 0.785 | -2.0 |

#### Binomial Regression Analysis (Table 4)

The Binomial regression model exhibits catastrophic failure across all countries, with universally negative *R*^2^ values ranging from -0.471 (Tanzania) to -2.666 (Mozambique)1, indicating predictions worse than a naive mean baseline. The average *R*^2^ of -1.611 demonstrates fundamental model inadequacy for this prediction task2. Climate integration provides negligible improvement (0.13% MAE reduction, 0.31% *R*^2^ improvement)3, confirming that the model’s core limitations—likely stemming from inappropriate distributional assumptions for continuous prevalence rates cannot be overcome through additional features. The particularly poor performance in Mozambique (*R*^2^ = −2.652) and Uganda (*R*^2^ = −1.979) suggests severe model-data mismatch in high-variability contexts4. MAE values, which range from 14.0 (Tanzania) to 39.7 (Mozambique), are 3-4 times higher than advanced models, underscoring the critical importance of appropriate model architecture selection5. This serves as a baseline demonstrating why sophisticated spatiotemporal approaches are necessary for malaria forecasting.

**Table 4:** Performance Comparison: Climate Model vs Baseline Model (Binomial) across five countries.

| Country | Climate Model |  |  | Baseline Model (Binomial) |  |  |
| --- | --- | --- | --- | --- | --- | --- |
| | MAE | $R^2$ | RMSE | MAE | $R^2$ | RMSE |
| Congo | 30.205 | -1.218 | 40.639 | 30.198 | -1.217 | 40.629 |
| Tanzania | 14.049 | -0.475 | 24.406 | 14.043 | -0.471 | 24.372 |
| Nigeria | 30.634 | -1.732 | 38.393 | 30.688 | -1.740 | 38.449 |
| Mozambique | 39.652 | -2.652 | 46.454 | 39.737 | -2.666 | 46.545 |
| Uganda | 33.887 | -1.979 | 41.512 | 33.956 | -1.988 | 41.577 |
| **Average** | **29.285** | **-1.611** | **38.281** | **29.324** | **-1.616** | **38.314** |

#### ESTT (Enhanced Spatiotemporal Transformer) Analysis (Table 5)

ESTT demonstrates strong benefits from climate integration, though the effect is not uniform (shown in Figure 4). The climate-enhanced model achieves a 3.6% average MAE reduction (8.404 vs. 8.718), 1.3% *R*^2^ improvement (0.752 vs. 0.742), and 1.8% RMSE reduction (11.352 vs. 11.560).Congo exhibits the most dramatic improvement with a 22.2% MAE reduction (5.907 vs. 7.590) and 2.0% *R*^2^ gain, suggesting that transformer attention mechanisms effectively leverage climate variables in high-transmission settings. Nigeria also shows substantial gains (1.9% MAE reduction, 4.5% *R*^2^ improvement).However, the improvements are not universal. Tanzania (2.% MAE increase), Mozambique (0.5% MAE increase), and Uganda (1.0% MAE increase) all show a slight performance degradation with the added climate data, even as *R*^2^ values remained relatively stable. This may reflect regional complexities, like microclimates, not captured by the simple temperature-precipitation features.Despite this, the uniformly high *R*^2^ values (0.694-0.818) demonstrate ESTT’s strong performance overall, and this architecture appears the most suited to exploiting climate information.

**Table 5:** Performance Comparison: Climate Model vs Baseline Model (ESTT) across five countries.

| Country | Climate Model |  |  | Baseline Model (ESTT) |  |  |
| --- | --- | --- | --- | --- | --- | --- |
| | MAE | $R^2$ | RMSE | MAE | $R^2$ | RMSE |
| Congo | 5.907 | 0.818 | 8.976 | 7.590 | 0.802 | 9.382 |
| Tanzania | 7.978 | 0.771 | 11.063 | 7.808 | 0.780 | 10.841 |
| Nigeria | 10.045 | 0.718 | 13.001 | 10.240 | 0.687 | 13.692 |
| Mozambique | 9.455 | 0.757 | 12.378 | 9.403 | 0.755 | 12.431 |
| Uganda | 8.633 | 0.694 | 11.344 | 8.549 | 0.688 | 11.456 |
| **Average** | **8.404** | **0.752** | **11.352** | **8.718** | **0.742** | **11.560** |

**Figure 4:**
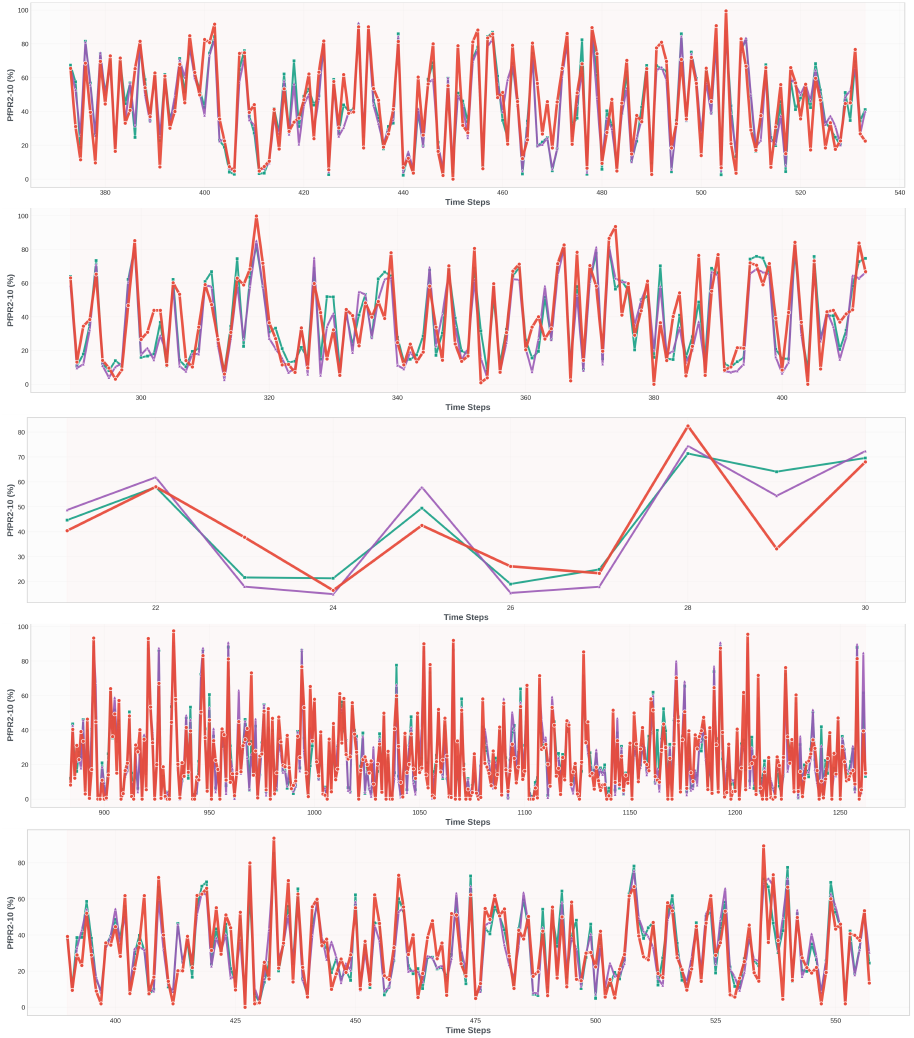
Future Forecast Details for Five Countries (Top to bottom: Mozambique, Nigeria, Congo, Tanzania, Uganda). Red: Actual values, Teal: Climate-enhanced model, Purple: Baseline model (ESTT)

#### PatchTST Analysis (Table 6)

PatchTST exhibits a negligible response to climate integration, with virtually identical performance between the climate and baseline variants. This remarkable stability across all countries suggests that the patch-based decomposition and channel-independent processing architecture may be inherently limited in leveraging cross-modal features like climate data. The model’s design—which processes each feature channel independently before aggregation—may prevent effective climate-epidemiological feature interactions that benefit transformer architectures with joint attention mechanisms.Performance is consistently weak across all regions (*R*^2^ = 0.063 – 0.365, MAE = 14.9-17.4), indicating fundamental limitations in capturing spatiotemporal malaria dynamics. Congo’s particularly poor performance (*R*^2^ = 0.063) suggests difficulty modeling high-variance equatorial transmission patterns. The marginal improvements in Nigeria (0.9% MAE reduction) and Tanzania (0.1% MAE increase) are well within noise margins. Overall, PatchTST’s patch-based approach appears ill-suited for this task, possibly due to: (1) loss of fine-grained temporal resolution through patching, (2) inability to model long-range dependencies critical for seasonal patterns, or (3) channel independence preventing climate-disease feature fusion. This architecture would require substantial modification to achieve competitive performance for malaria forecasting.

**Table 6:** Performance Comparison: Climate Model vs Baseline Model (PatchTST) across five countries.

| Country | Climate Model |  |  | Baseline Model (PatchTST) |  |  |
| --- | --- | --- | --- | --- | --- | --- |
| | MAE | $R^2$ | RMSE | MAE | $R^2$ | RMSE |
| Congo | 16.221 | 0.063 | 20.389 | 16.096 | 0.068 | 20.333 |
| Tanzania | 14.984 | 0.310 | 19.210 | 14.962 | 0.305 | 19.287 |
| Nigeria | 17.289 | 0.262 | 21.019 | 17.446 | 0.257 | 21.097 |
| Mozambique | 16.370 | 0.365 | 20.013 | 16.296 | 0.363 | 20.044 |
| Uganda | 14.909 | 0.174 | 18.644 | 14.979 | 0.176 | 18.615 |
| **Average** | **15.955** | **0.235** | **19.855** | **15.956** | **0.234** | **19.875** |

#### Cross-Model Analysis

Our comparative analysis across the different architectures reveals several important findings. The **ESTT** model achieved the best overall performance, with an average MAE of 8.404 and an *R*^2^ of 0.752, and it consistently improved when climate data was included. This validates the effectiveness of its attention-based mechanisms for integrating spatiotemporal information. The **Neural ODE** also proved to be a strong baseline, achieving an MAE of 9.473 and an *R*^2^ of 0.734. Surprisingly, its performance slightly worsened with added climate features, suggesting its continuous temporal dynamics might already capture the underlying climate-driven patterns. Traditional ensemble methods like **Random Forest** and **Gradient Boosting** were also competitive, with *R*^2^ values ranging from 0.7 to 0.8, although their response to climate data varied significantly by region.

In sharp contrast, some modern architectures were not well-suited for this task. **PatchTST** consistently underperformed across all countries (MAE ≈ 16, *R*^2^ ≈ 0.23), indicating its patch-based design is a poor fit. **Binomial Regression** failed entirely, producing negative *R*^2^ values and confirming that advanced spatiotemporal models are necessary. This highlights a substantial performance gap, with the weaker models showing error rates 2-3 times higher than top performers. Ultimately, the impact of climate data is highly dependent on the model’s architecture: transformer-based models like ESTT thrive on the extra data, while others like Neural ODE and PatchTST see minimal or even negative effects, showing that the model’s design is key to utilizing environmental information.

#### Uncertainty Estimation Analysis (Table 7)

Our comparative analysis reveals several important findings (shown in Figure 5). The ESTT model achieved the best overall average performance (MAE 8.404, *R*^2^ 0.752), validating its strong architecture. However, its response to climate data was mixed: while the average performance improved, the model’s accuracy actually worsened in 3 of the 5 countries, suggesting its benefits are region-specific.The Neural ODE proved to be a strong baseline (MAE 9.473, *R*^2^ 0.734), but as noted, its performance slightly worsened with added climate features. Traditional ensemble methods like

**Table 7:** Uncertainty Quantification Results Across Models.

| Metric | MC-PatchTST | MC-ESTT | MC-Neural ODE |
| --- | --- | --- | --- |
| **Prediction Uncertainty ($\sigma$)** |  |  |  |
| Mean | 3.7388 | 2.7302 | 3.8210 |
| Median | 3.1900 | 2.1873 | 3.7062 |
| **68% Prediction Intervals** |  |  |  |
| Coverage (%) | 23.4 | 38.6 | 22.1 |
| Avg. Width | 7.4775 | 5.4604 | 7.6420 |
| **95% Prediction Intervals** |  |  |  |
| Coverage (%) | 42.1 | 57.0 | 46.3 |
| Avg. Width | 14.6559 | 10.7023 | 14.9783 |

**Figure 5:**
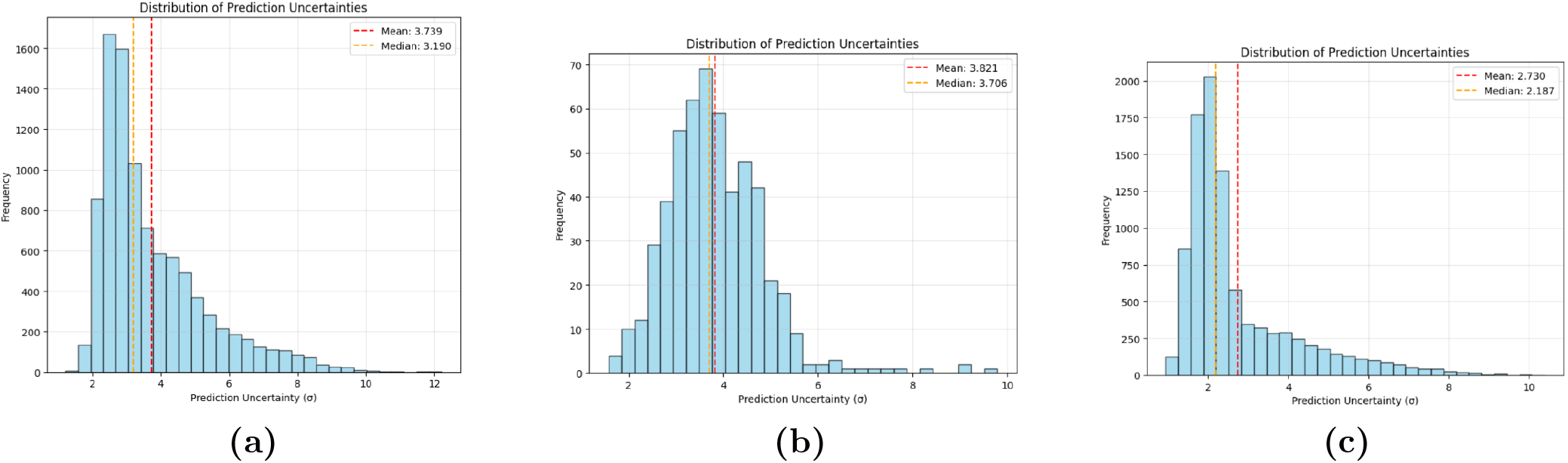
Uncertainty Quantification Analysis: **(a)** UA-PatchTST, **(b)** UA-Neural ODE, **(c)** UA-ESTT.

Random Forest and Gradient Boosting were also competitive, with *R*^2^ values ranging from 0.626 to 0.805, though their response to climate data varied significantly by region.In sharp contrast, some architectures were not well-suited for this task. PatchTST consistently underperformed (MAE ≈ 16, *R*^2^ ≈ 0.23), and Binomial Regression failed entirely with negative *R*^*2*^ values. This highlights a substantial performance gap, with weaker models showing error rates 2-3 times higher than top performers. Ultimately, the impact of climate data is highly dependent on the model’s architecture. While the ESTT’s average performance benefited, others like Neural ODE and PatchTST saw minimal or negative effects, showing that model design is key to utilizing environmental information.

## 6 Discussion and Future Work

Our findings demonstrate that deep learning models, specifically the Enhanced Spatiotemporal Transformer (ESTT) and Neural ODE, significantly advance malaria prediction, achieving error rates 2-3 times lower than traditional approaches. The ESTT model yielded the highest accuracy (MAE = 8.404, *R*^2^ = 0.752) and also excelled at uncertainty quantification, achieving the most dependable and best-calibrated intervals. The Neural ODE provided competitive performance (MAE = 9.473, *R*^2^ = 0.734), but its uncertainty estimates showed poor calibration. This ability to provide confidence-aware forecasts is critical for risk-based public health planning. Our analysis also revealed that model architecture dictates the utility of climate data; transformer-based models benefited on average, whereas the Neural ODE’s continuous-time dynamics appeared to inherently capture these seasonal patterns, making the explicit features redundant. The poor performance of PatchTST (MAE ≈ 16, *R*^2^ ≈ 0.23) further confirms that a specialized architecture is essential for this complex task.

A deployment-ready pipeline can also be developed by wrapping the best-performing model into a lightweight API (e.g., using FastAPI) and containerizing it with Docker for ease of integration into public-health dashboards. Such a system would allow health agencies to upload new climate or surveillance data and obtain real-time malaria prevalence forecasts with associated uncertainty estimates. This potential deployment pathway would make the proposed models practically usable for operational disease monitoring and decision support.

## Data Availability

All data produced are available online at http://dx.doi.org/10.7910/DVN/Z29FR0
Climate data can be provided upon reasonable request to the authors.

## Author Contributions

The first two authors (Vikalp and Manoj) conceived the study and designed the modeling framework. They also implemented the deep learning models, conducted the experiments, and performed the data analysis. Sanika contributed to the statistical modeling. All authors jointly interpreted the results and guided methodological choices. Vikalp, Manoj, and Abdessamad prepared the initial manuscript draft, and all authors contributed to revisions and approved the final version.

## Competing interest

The authors declare no competing interests.

## Funding Statement

This research received no external funding.

## A Appendix

In this section, we provide more details about Enhanced Spatio-Temporal Transformer and PatchTST based Spatio-Temporal Forecasting Architectures.

### A.1 Nonlinear Spatiotemporal Operator Interpretation

We model malaria prevalence forecasting as learning a nonlinear spatiotemporal operator:

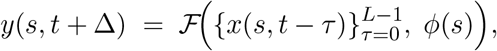

where 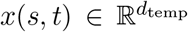 denotes temporal covariates (e.g., rainfall, temperature, previous prevalence) at location *s* and time *t*, and 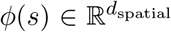 denotes static spatial descriptors (e.g., geographic coordinates, elevation, population density). The operator ℱ is parameterized by the proposed enhanced spatiotemporal transformer architecture.

#### A.1.1 Temporal Transformer as Data-Adaptive Temporal Convolution

In each self-attention block, given temporal embeddings 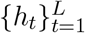 :

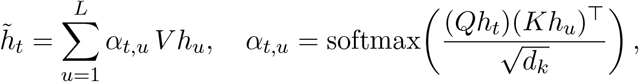

where *Q, K, V* are learned projections. Interpreting *α*_*t,·*_ as a *data-dependent kernel K*_*θ*_(*t, u*; **x**), we have:

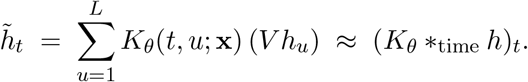

Thus, multi-head attention performs a **nonlinear, input-conditioned convolution** over the temporal dimension. Stacking layers with feedforward networks yields a *Volterra-type operator*, which is well-suited for capturing delayed rainfall–vector–transmission pathways in malaria.

#### A.1.2 Spatial MLP as Location Embedding

Spatial features are mapped via:

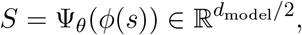

where Ψ_*θ*_ is a multi-layer perceptron with normalization, GELU activation, and dropout. This embedding encodes *static modifiers* such as endemicity, ecological factors, and connectivity.

#### A.1.3 Temporal–Spatial Fusion as Cross-Term Interaction

Fused representations are obtained via attention:

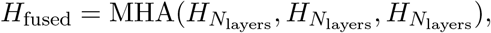

followed by temporal pooling:

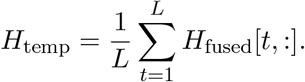

The final prediction is:

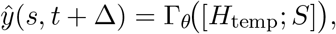

where Γ_*θ*_ is the prediction MLP. Since *H*_temp_ already incorporates attention weights influenced by *ϕ*(*s*) through shared parameters, this effectively learns:

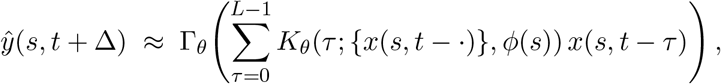

a **nonlinear spatiotemporal convolution** with spatially modulated, history-dependent kernels.

#### A.1.4 Approximation Capacity

Transformers with GELU activations and MLPs are universal sequence-to-scalar approximators on compact sets. Hence, the architecture can approximate continuous operators such as Volterra or Wiener functionals, which capture lagged climate effects in malaria dynamics. Learnable positional embeddings ensure temporal order awareness, preventing permutation invariance in time.

#### A.1.5 Stability Properties

With norm-first residual blocks, the per-layer Lipschitz constant satisfies:

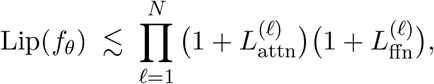

where 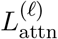 and 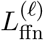 are layer-specific constants. Layer normalization keeps these factors moderate, while dropout reduces overfitting. Temporal averaging (pooling) is a contractive step (operator norm ≤ 1), further stabilizing predictions.

#### A.1.6 Domain Alignment

Malaria prevalence depends on multi-month, nonlinear lags (rainfall → mosquito breeding → transmission). Data-adaptive kernels capture *when* and *how strongly* to weight past months. Spatial embeddings inject location-specific priors, allowing kernel shapes to vary across regions.

### A.2 Patch-Based Time Series Decomposition

The core insight of PatchTST is that time series can be effectively decomposed into non-overlapping or overlapping patches that capture local temporal patterns more efficiently than full-sequence attention mechanisms.

#### A.2.1 Patch Creation and Embedding

For a given time series *X* ∈ ℝ^*T ×d*^, we create patches of length *P* with stride *S*:

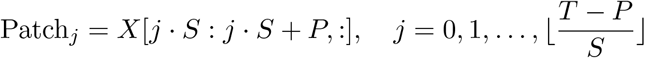

Each patch is then flattened and linearly projected to the model dimension:

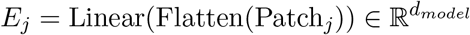

where the linear projection transforms patches from ℝ^*P ×d*^ to 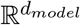. This patch-based approach reduces the sequence length from *T* to 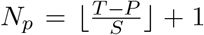 patches, significantly improving computational efficiency while preserving local temporal structure.

#### A.2.2 Positional Encoding for Patches

To maintain temporal ordering information, we add learnable positional encodings to patch embeddings:

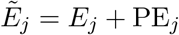

where 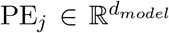 represents learnable position embeddings that encode the temporal position of each patch within the sequence.

### A.3 Channel-Independent Processing Architecture

A fundamental innovation of PatchTST is channel-independent processing, where each feature dimension is processed by a separate transformer encoder. This approach is particularly beneficial for malaria forecasting where epidemiological variables (age groups, examination counts) and climate variables (temperature, precipitation) exhibit different temporal dynamics and scaling properties.

#### A.3.1 Individual Channel Transformers

For each feature channel *c* ∈ {1, 2, …, *d*}, we extract the corresponding data:

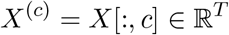

Channel-specific patches are created and processed by dedicated transformer encoders:

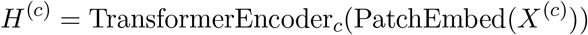

where each TransformerEncoder_*c*_ consists of *L* transformer layers with multi-head self-attention:

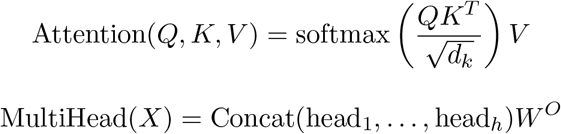

where 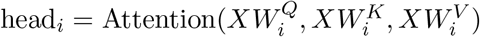.

#### A.3.2 Channel Aggregation and Global Representation

After independent processing, channel representations are aggregated to form a unified temporal representation:

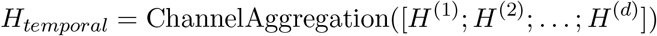

where [·; ·] denotes concatenation and the aggregation function learns optimal channel combination:

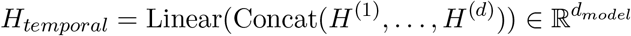

### A.4 Spatial Feature Integration

To account for the significant spatial heterogeneity in geographic and climatic conditions, our architecture explicitly models location-specific attributes. This is achieved through a dedicated sub-network that processes geospatial features to learn a distinct embedding for each location, thereby conditioning the forecast on its unique regional context.

#### A.4.1 Spatial Feature Projection

Spatial features 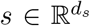 including coordinates and location-specific climate statistics undergo projection and nonlinear transformation:

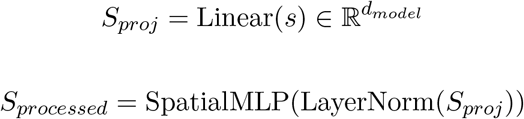

where the spatial MLP consists of multiple layers with GELU activations and dropout regularization:

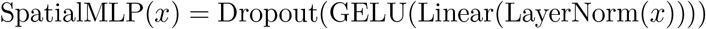

### A.5 Attention-Based Spatiotemporal Fusion

The integration of temporal and spatial representations employs cross-attention mechanisms that learn optimal feature interactions for malaria prediction.

#### A.5.1 Cross-Attention Fusion

Temporal features serve as queries while spatial features provide additional context through key-value pairs:

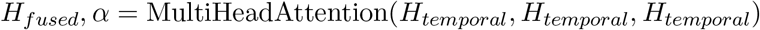

where the attention weights *α* provide interpretability regarding which temporal periods contribute most to predictions.

#### A.5.2 Final Prediction Network

The fused representation combines temporal and spatial information for prevalence prediction:

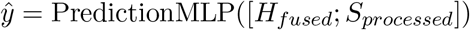

The prediction network employs multiple layers with residual connections and layer normalization:

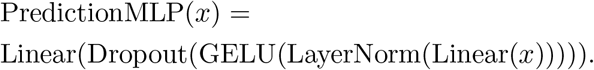

### A.6 Training Methodology and Optimization

#### A.6.1 Loss Function and Regularization

Training employs Smooth L1 Loss for robustness against outliers common in epidemiological data:

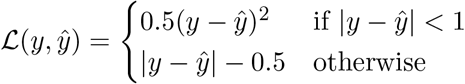

L2 regularization is applied to prevent overfitting:

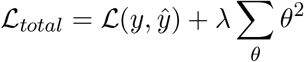

#### A.6.2 Advanced Optimization Strategy

The optimization employs AdamW with cosine annealing warm restarts to escape local minima:

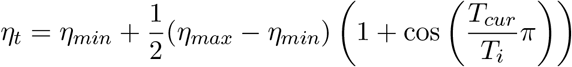

where *T*_*cur*_ represents the current epoch within restart cycle *T*_*i*_, and 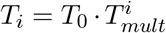 grows exponentially to allow for longer exploration periods.

Gradient clipping with maximum norm constraint ensures training stability:

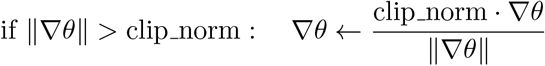

## Notes

### Competing Interest Statement

The authors have declared no competing interest.

### Funding Statement

This study did not receive any funding.

### Author Declarations

Data is available from Harvard Dataverse, V1, under a CC-BY 4.0 license. Source - http://dx.doi.org/10.7910/DVN/Z29FR0

### Summary of Updates

Updated author details. Added authors contribution details. Funding statement and competing interest statement is also added.

